# Competing Risk Survival analysis of time to in-hospital mortality or Recovery among Covid-19 Patients in South-East Ethiopia: a hospital-based multisite study

**DOI:** 10.1101/2024.06.04.24308446

**Authors:** Addis Wordofa, Ayalneh Demissie, Abdurehman Kalu, Abdurehman Tune, Mohammed Suleiman, Abay Kibret, Zerihun Abera, Yonas Mulugeta

**Affiliations:** Department of Public Health, College of Health Science, Arsi University, Ethiopia; Department of Anesthesia, College of Health Science, Arsi University, Ethiopia; Department of Nursing, College of Health Science, Arsi University, Ethiopia; Department of Biomedical, College of Health Science, Arsi University, Ethiopia

**Author notes:** **Corresponding Author**: Ayalneh Demissie. AW: Addis Wordofa, AD: Ayalneh Demissie, AKa: Abdurehman Kalu, AT: Abdurahman Tune, MS: Mohammed Suleiman, AKi: Abay Kibret, ZA: Zerihun Abera, YM: Yonas Mulugeta.

**Keywords:** Time to death, Recovery, Censoring, COVID- 19, Competing risks, Survival analysis, Ethiopia

## Abstract

**Background:** To date, survival data on risk factors for COVID-19 mortality in south- Ethiopia is limited, and none of the published survival studies have used a competing risk approach. This study aims to identify risk factors for in-hospital mortality in COVID-19 patients hospitalized at one of the six hospitals in southeast -Ethiopia, considering recovery as a competing risk.

**Methods:** This observational multisite study included a medical record of 827 confirmed SARS-CoV-2 cases hospitalized at one of the six hospitals in southeast-Ethiopia from October 1, 2022 to May 31, 2023. We compiled data on the patients’ socio-demographic characteristics, clinical manifestation, comorbidity, treatment status, treatment outcomes, and length of stay. We performed a Cox regression analysis for competing risks, presenting cause-specific hazard ratios (HRcs) for the effect of preselected factors on the absolute risk of death and recovery.

**Results:** 827 patients were included (51.9% male; median age 50 years, IQR: 38—65). Patients were hospitalized for a median duration of 5 days (IQR: 1—7); 139 (17%) of them died, while 516 (62%) were recovered and discharged alive, the rest 172 (21%) were censored. Patients with higher age (HRcs 2.62, 95% CI 1.29—5.29), immune-compromised state (HRcs 1.46, 95% CI 1.08—1.98) had increased risk of death, whereas male sex paradoxically (HRcs 0.45, 95% CI 0.22—0.91) associated with decreased risk of death. We found no increased mortality risk in diabetes patients.

**Conclusion:** This competing risk survival analysis allows us to corroborate specific pattern of risk factors about COVID-19 mortality and its progression among different groups of individuals (differentiated by age and immune-compromised state). 62% presenting cases recovered within a median duration of 5 days; where as 17% die within the first 72 hours, most with immune-compromised conditions. This should be considered while planning and allocating the distribution of care services for effective health service delivery

## Introduction

Despite limited access to healthcare [1,2] and relatively milder social distancing restrictions compared to those imposed in most high-income countries [3,4], corona virus disease 2019 (COVID-19) mortality rates have been relatively low throughout Africa [5]. As of June 23, 2021, the World Health Organization (WHO) reports 3,852,707diagnosed cases and 92,719 deaths in the continent [5]. However, severe acute respiratory syndrome corona virus 2 (SARS-CoV-2) transmission and survival dynamics have been highly heterogeneous across different African countries in terms of timing and implemented interventions [6].

In sub-Saharan Africa, Ethiopia is second only to South Africa in terms of the number of recorded cases and deaths, with an overall case fatality ratio (CFR) of about 1.5% compared to about 2.2% in the rest of the world [5]. The first COVID-19 case was confirmed on March 13, 2020, meanwhile, the Ethiopian government declared a state of emergency on April 8, 2020 in the country [7]. Since then, rigorous contact tracing, isolation, and compulsory quarantine have been established [8,9]. Schools and borders closed, public institutions and firms operated at minimum capacity or closed completely, and people informed to stay at home [8]. Nevertheless, in November 2020, schools reopened in the entire country, and social gatherings up to 50 individuals were allowed again. As of June 23, 2021, 275,391 Covid-19 cases and 4290 deaths [5] were recorded nationally, with thousands of cases reported in all the 12 regions of Ethiopia [9]. In Ethiopia, a syndromic surveillance is carried out to identify Covid-19 infected individuals.

Samples from suspected cases and case contacts are collected at different health facilities displaced in the country (including health centers serving the most rural areas) and cases are confirmed via real-time reverse transcription–polymerase chain reaction (RT-PCR) test.

Collected samples are analyzed by 38 national, regional, hospital, and private laboratories [10]. Both suspected and confirmed cases are admitted to isolation centers and discharged after a negative laboratory test [9]. Although swab testing was initially applied to both symptomatic patients and all close contacts of cases, it is possible that, due to limited resources and the increased number of cases in the country, only symptomatic case contacts are currently tested. Active monitoring of cases conducted by the Ethiopian Public Health Institute suggested that 52% of the identified positive cases were asymptomatic [11]. As of January 10, 2021, the overall rate for positive laboratory test results since the first detection of the epidemic in the country was 6.9%, likewise 1,054 and 524 Intensive Care Unit(ICU) beds and mechanical ventilators in the COVID-19 treatment centers for an estimated population of 117million [11].

The possible spread of COVID-19 in rural areas of the country is especially dangerous because of the sparse presence of well-resourced health facilities implying long travel distances for remote populations, which is an important barrier to universal access to primary care[2].

Moreover, the healthcare workforce in Ethiopia is 5 times lower than the minimum threshold defined by the WHO for Sustainable Development Goals health targets [12] and far below the African average [13].

Unfortunately, new facts come with a lag compared to the virus spread and governments are forced to make prompt decisions based on limited evidence which changes at a staggering pace.

The fight against a practically unknown enemy has been and still is the major obstacle. Aside from studying the virus’s biology, infecting mechanisms, probable treatments and of course vaccine development, epidemic modeling has stepped forward.

In a trade-off between recovery and death, competing risk modeling strategies provide a sharp suit that allows exploring a variety of scenarios and provides an intuitive understanding of the most critical factors governing disease dynamics.

Recent modeling studies have made a difference for public health care decision making by providing, for example, estimations of the impact of Non-Pharmaceutical Interventions (NPI) in a number of sub-Saharan African countries, highlighting the difficulties in defining effective, feasible, and sustainable strategies for suppression or mitigation of COVID-19 epidemics [14–17].

The main challenge is to create a model that predicts plausible scenarios for a disease we have known for only a few months. One of the most important barriers for the provision of solid epidemiological parameters has been the different management strategies that each country has taken in response to this outbreak.

Variation in survival cannot be explained only by the different population age structure, symptom data, specific clinical parameters or available critical care beds.

Because of the increase in mortality due to COVID-19 and the increase in the speed of its spread, many methods have been developed to reliably predict patient survival based on symptom data and specific clinical parameters. Studies describing the clinical features of COVID-19 and risk factors associated with incidence and timing of poor outcome have been extensively published [18], but survival data regarding risk factors for COVID-19 mortality in south-east Ethiopia is limited. Further- more, to our knowledge, none of the published survival studies have considered recovery as competing risk for mortality. Not taking competing risks into account leads to biased mortality estimates and to overestimation of survival curves [19,20]. Analyzing mortality data with a more accurate competing risk analysis adds to the growing body of evidence on disease course and risk factors of a poor COVID-19 outcome. More robust knowledge about these risk factors is crucial to inform local prediction research [18]. Calculating the probability to survive and the effect of each feature like symptoms (competing risks) in our case on survival probability was done using survival analysis. Survival analysis is a model for time until a certain “event”. Time-to-event data encounters several research challenges such as censoring, symptoms correlations, high-dimensionality, temporal dependencies, and difficulty in acquiring sufficient event data in a reasonable period of time [21]. Most importantly not taking, competing risks into account leads to biased mortality estimates and to overestimation of survival curves. Therefore, evidence showing the duration of recovery and pace of death from COVID-19 in different contexts and settings is necessary for tailoring appropriate treatment and prevention measures.

There are many current literature techniques for conducting this sort of survival study. Among them, the Fine and Gray model have been deployed for computing infection risks, performing survival analysis and classification [22]. Therefore, the above model should be able to take these variables into account.

The present study aimed to analyze the socio-demographic and clinical profile of COVID-19 patients that were hospitalized to one of the six hospitals in the southeast-Ethiopia, characterized by different levels of access to healthcare while accounting for the effect of gender, location, clinical presentation, and co-morbid conditions on mortality and recovery.

## Methods

### Study setting

The study was conducted at six hospitals in southeast Ethiopia: Bishoftu, Modjo, Adama, Negele Arsi, Bekoji and Shashamane hospitals, also COVID-19 isolation and treatment Centers located in southeast Ethiopia, 35, 75,85,229,232 and 270kms from Addis Ababa (the capital), respectively. The isolation and treatment centers were the place where all Ethiopian and none-Ethiopian citizens with COVID-19 were admitted for isolation, care and support. There are 82 functional public hospitals in the region of which 44 are primary hospitals and 34 are general hospitals and 4 are comprehensive specialized hospitals [23].

### Study design

We performed an observational multisite study using a medical record of 827 confirmed SARS-CoV-2 cases hospitalized at one of the six sites: Bishoftu, Modjo, Adama, Negele Arsi, Bekoji and Shashamane, also called COVID-19 treatment centers from October 1, 2022 to May 31, 2023. The sample size for duration of recovery was calculated taking the following assumptions into account: 95% Confidence Interval (CI), 46% survival probability, 5% margin of error and 10% withdrawal, and equal proportion of individuals in each group [24]. The minimum sample size required to conduct this study was 419 with a design effect 2 due to differences in the size of the hospitals, the final sample size become 838.

### Sampling procedures

To select the records studied, we consecutively included confirmed adult patients record with COVIC-19, and who were admitted to one of the six hospitals in Oromia region from Bishoftu to Shashemene for at least 24 hours between October1, 2022 and May 31, 2023. In the current analysis, we excluded patients if data regarding duration of hospital admission were missing for those who died or recovered. A sample of 827 patients allocated proportional to their catchment population size (PPs) to each center = n_x_*n/N

|  |  |  |  |
| --- | --- | --- | --- |
| 1. Bishoftu (KTC) | $n_1=750$ | final $n_{1f}$ | 171 |
| $\frac{750}{3631} * 827 = 171$ | | | |
| 2. Modjo PH | $n_2=605$ | final $n_{2f}$ | 138 |
| $\frac{605}{3631} * 827 = 138$ | | | |
| 3. Adama (AHMC) | $n_3=931$ | final $n_{3f}$ | 212 |
| $\frac{931}{3631} * 827 = 212$ | | | |
| 4. Bekoji PH | $n_4=650$ | final $n_{4f}$ | 148 |
| $\frac{650}{3631} * 827 = 148$ | | | |
| 5. Shashamane RH | $n_5=396$ | final $n_{5f}$ | 90 |
| $\frac{396}{3631} * 827 = 90$ | | | |
| 6. Negele Arsi PH | $n_6=299$ | final $n_{6f}$ | 68 |
| $\frac{299}{3631} * 827 = 68$ | | | |
| Total | <u>$N=3631$</u> |  | <u>$n= 827$</u> |

### Variables collected and definitions

#### Outcome variables (dependent variables)

The possible outcomes categorized as a recovered case or death. The primary outcome was COVID-19 death that reported as a death occurring in a confirmed COVID-19 case. Recovery/ survival were analyzed as competing event/ secondary endpoint. COVID-19 patients who were discharged due to clinical improvement, for medical rehabilitation, or transfer to a nursing home were considered ‘recovered’. COVID-19 patients who were transferred to a non-study hospital, who were lost to follow-up because of other reasons, or whose reason of discharge is unknown or missing were considered ‘censored ’.

#### Covariates (independent variables)

The socio-demographic factors, health-related factors, co-morbidities, clinical manifestation, laboratory -result and treatment-related factors were considered independent variables.

Outcome variables: The possible outcomes categorized as a recovered case or death. The primary outcome was in-hospital mortality. Recovery/ survival were analyzed as competing event/ secondary endpoint. COVID-19 patients who were discharged due to clinical improvement, for medical rehabilitation, or transfer to a nursing home were considered ‘recovered’. COVID-19 patients who were transferred to a non-study hospital, who were lost to follow-up because of other reasons, or whose reason of discharge is unknown or missing were considered ‘censored ’.

### Data collection

We extracted routine data of all confirmed COVID-19 patients that were followed from admission to one of the six treatment sites till discharge using a structured datasheet. It includes socio-demographic characteristics, status during admission, information on clinical presentation, diagnostic procedure, comorbidity, type of co-morbidity, disease course, treatments, type of drug used, treatment outcomes and length of stay. Five public health officers collected the data after we provided them two-days training on the data collection instruments. Patient data were entered anonymously into electronic case form (using Epidata Entry client), only using a study identifier. The middle six authors (AD, AKa, AT, MS, AKi and ZA) supervised the data collection and entry for completeness in real-time.

### Statistical analysis

Collected data were coded, entered, cleaned and analyzed using STATA software Version 16.1. Descriptive summary statistics such as: mean, standard deviations (SD) and inter quartile range (IQR) were calculated for continuous variables, while frequencies, and percentages for categorical variables. Moreover, we used a competing risks regression model to evaluate the effect of risk factors for time-to-event analyses or on the time from admission to death and on the time from admission to recovery [25,26]. A study evidenced the occurrence of competing risk if subjects experience one or more events or outcomes which compete with the outcome of interest [26]. In our study, recovery is considered a competing risk for mortality and is taken into account as an extra outcome, whereas in standard survival analyses, patients who recover are censored. However, the latter violates the assumption of non informative censoring, i.e. the recovered patients are not representative of those who are still admitted to the hospital in terms of their risk of dying. Censoring recovered patients induces bias and overestimation of survival curves, i.e. Kaplan Meier estimate incidence of death with upwards biases [25,26]. For the competing risk analysis, we estimated univariable and multivariable cause-specific hazard ratios (HRcs) for death and recovery for selected risk factors [20, 25]. These risk factors were pre-selected based on literature and expert opinion to be clinically relevant and routinely available at time of presentation, rather than based on statistical significance [27]. Variables with a P-value of less than or equal to 0.25 were considered as candidates for multivariate analysis. Variables in the final model with a p-value < 0.05 were considered statistically significant. The results were expressed as adjusted hazard ratios (HR) and their 95% confidence intervals (CI). Furthermore, cumulative incidence probabilities were estimated using the Fine and Gray approach [20, 25]. Gray’s test was used to compare equality of cumulative incidence curves (CIFs) across subgroups [28,29]. The proportional hazards assumption was checked by an evaluation of the Schoenfeld residuals.

Multivariable HRcs were also estimated for a subpopulation of patients with a ‘non ICU admission ’. The cause-specific hazard (CSH) model was fitted again for this subpopulation, to assess the influence of risk factors on the outcome in these patients. Variables with a P-value of less than or equal to 0.25 were considered as candidates for multivariate analysis. Variables with a P-value of less than 0.05 were considered statistically significant.

### Missing data

Missing data in the variables ‘age, gender, religion, smoking status, clinical manifestation, type of co morbidity, status at admission, dexamethasone use and length of stay were assumed to be missing at random (MAR) and the missed count were dropped from the analysis in these nine datasets.

## Results

A total of 827 confirmed COVID-19 patient records were included for analysis, the rest 11 patient records that had fragmentary data were excluded. Among them, 516 (62%) patients were recovered, 172 (21%) were censored and the remaining 139 (17%) patients were dead of COVID-19 in the hospital.

The median follow-up time were 17.7 (95% CI: 14.5–21.4) days. The cumulative incidence of both death and recovery increased over time. The median duration of hospital admission until death and recovery was 5 days (IQR: 1—7) and 11 days (IQR: 5—16), respectively. The median age of patients were 50years (IQR: 58—77), most 429 (51.9%), 601(72.7%) and 637 (77%) were male, from urban and presented with clinical manifestations, respectively. Nearly half patients 384(46.4%) had one or more co morbidities as shown in Table 1.

**Table 1.**
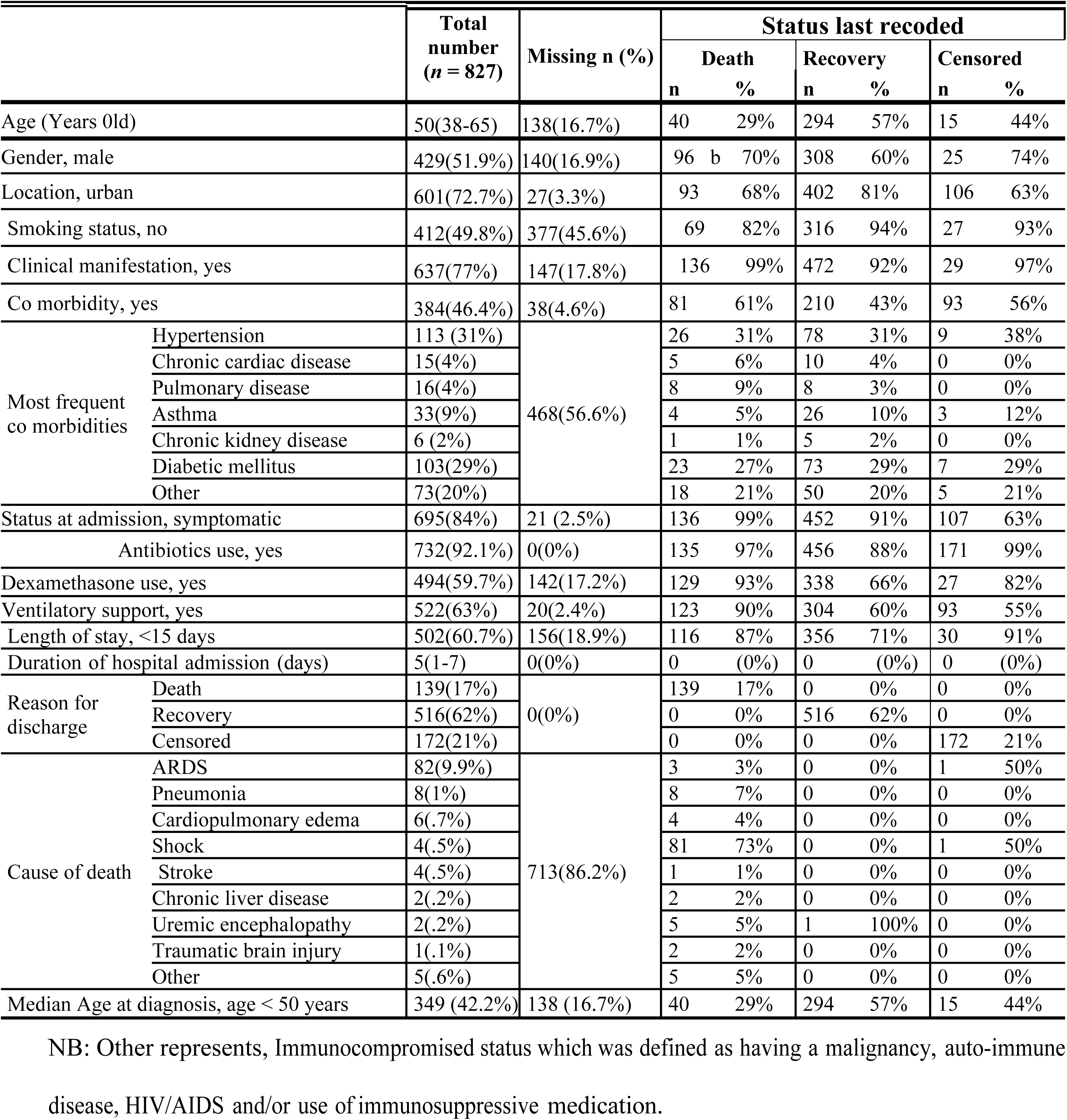
Characteristics of the study population in hospitals, south eastern Ethiopia 2024.

### Cumulative incidence curves

The Cumulative Incidence function (CIF) curves of the total patients showed that the probability of death after one, two and three weeks of hospital admission was 22.3% (95% 15.2—32.0), 35.1% (95% CI 23.7—49.9), and 51.3% (95% CI 32.1—73.9), respectively (Fig 1).

**Figure 1.**
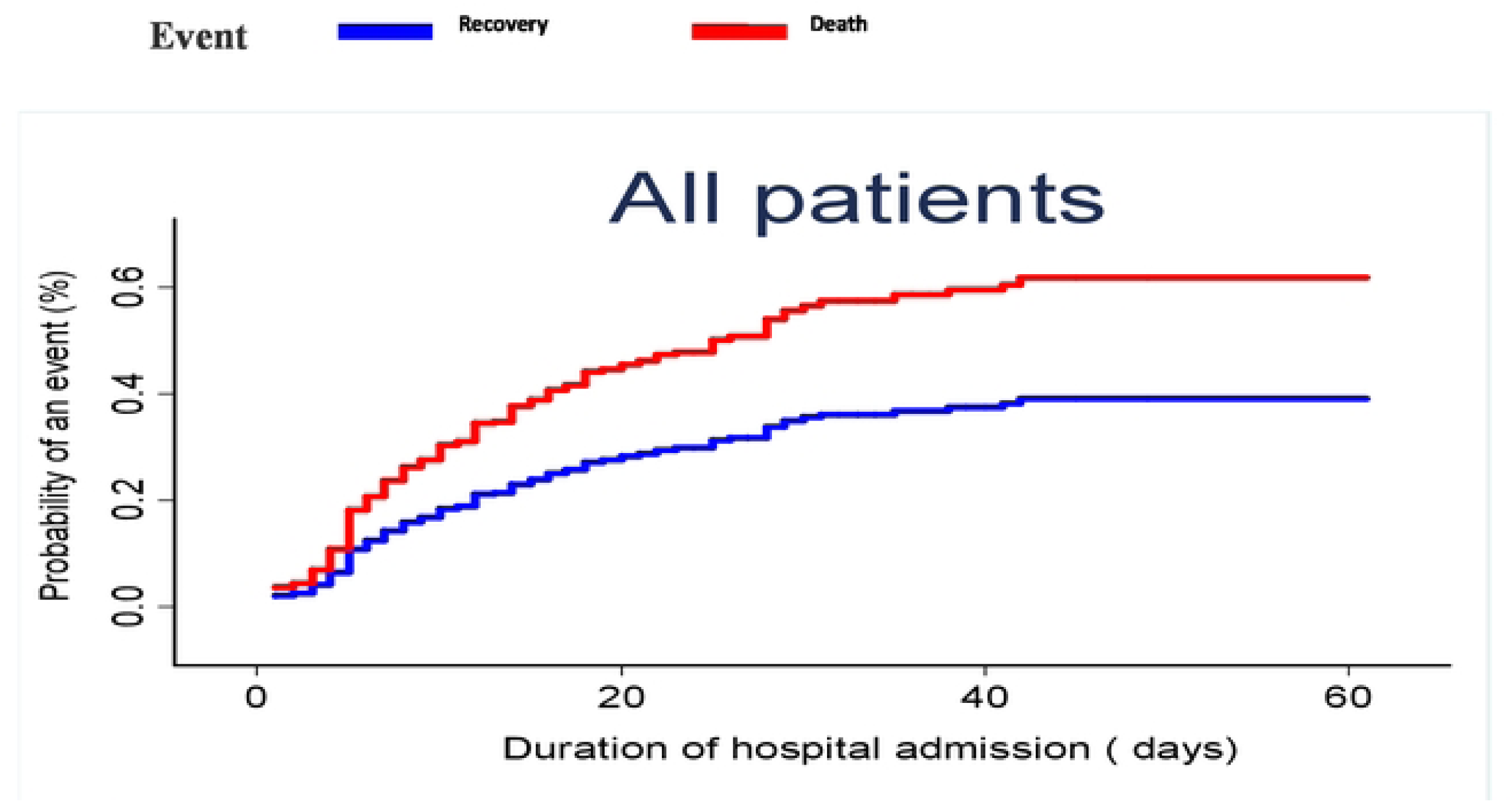
Cumulative incidence plot of death and recovery in south eastern Ethiopia hospitals, 2024.

The probabilities of recovery were 7.6% (95% 5.6—10.3), 17.9% (95% CI 14.3—22.2), and 44.7% (95% CI 36.2—54.1), respectively. Patients aged ≥ 50 years had a higher chance of death (p<0.01) and a lower chance of recovery (p<0.05) than patients aged <50 years (Fig 2). Males had a lower probability of recovery and a higher probability of death (p = 0.004 and p = 0.003) than females (Fig 3).

**Figure 2.**
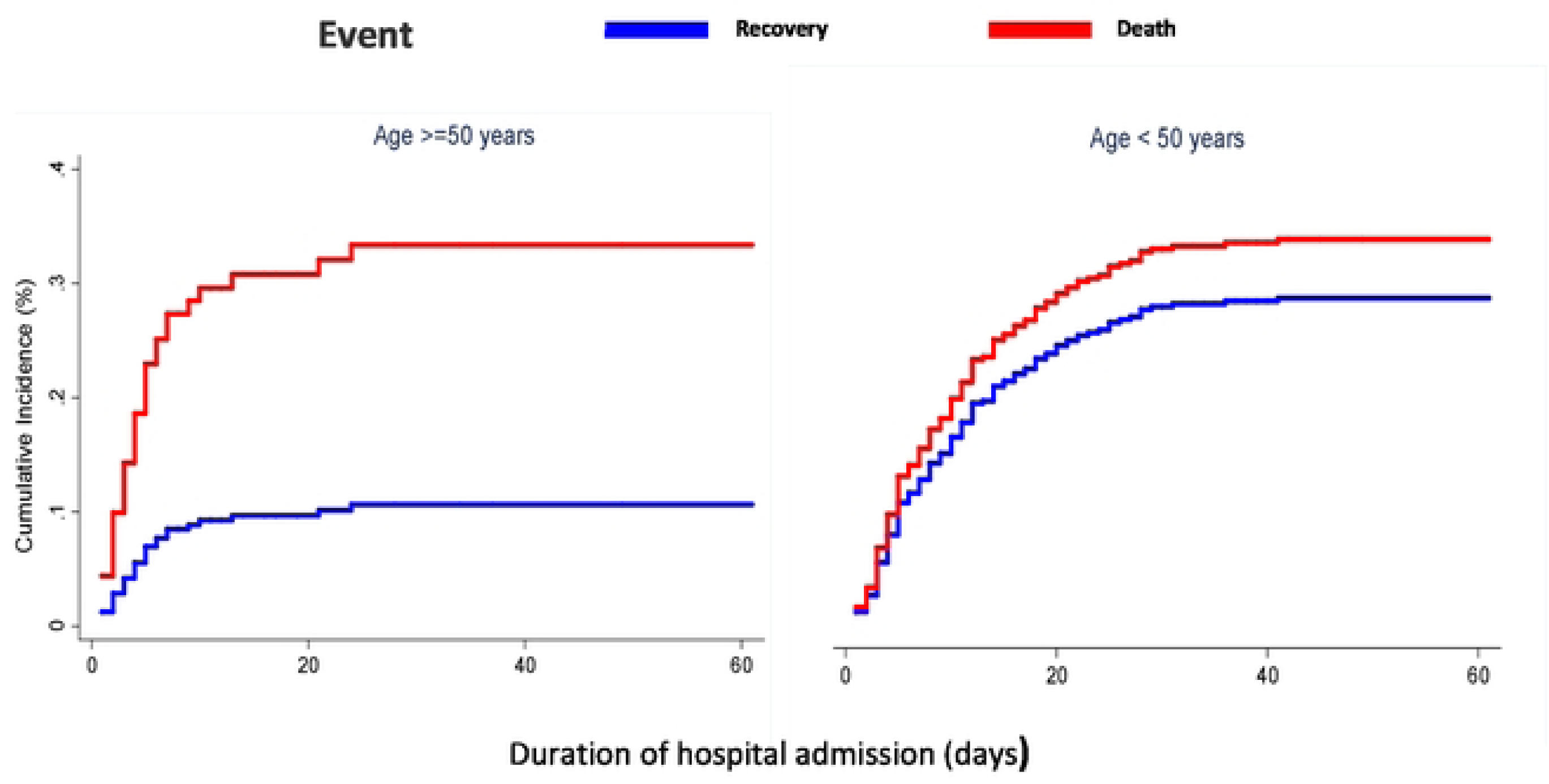
Cumulative incidence plots of death and recovery in south eastern Ethiopia hospitals, separated by age group 2024.

**Figure 3.**
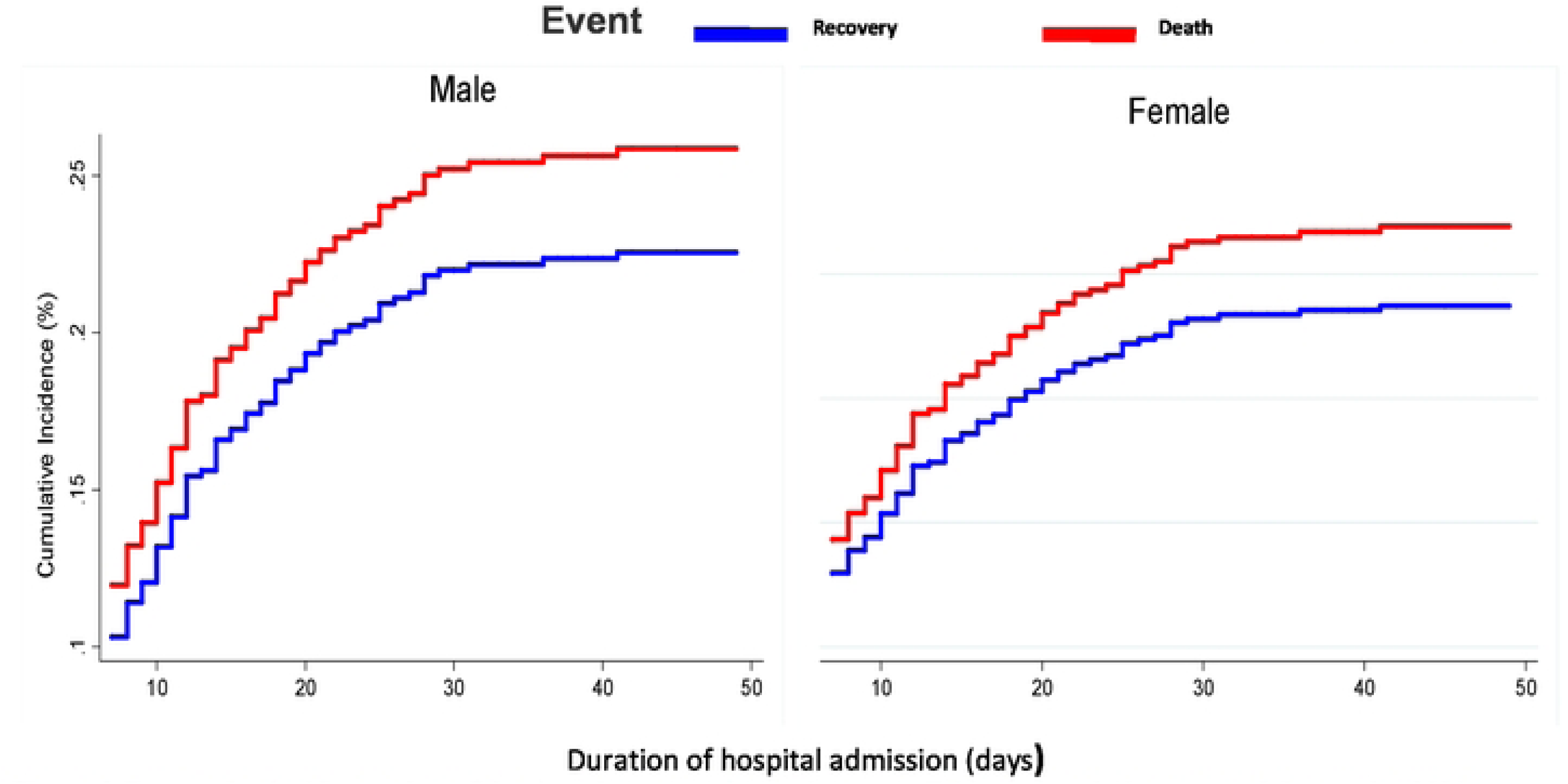
Cumulative incidence plots of death and recovery in south eastern Ethiopia hospitals, separated by gender 2024.

As shown in figure 1 cumulative incidence plot of death and recovery in the total patients, the probability of death conditional on not having recovered after one, two and three weeks of hospital admission was 22.3% (95% 15.2—32.0), 35.1% (95% CI 23.7—49.9), and 51.3% (95% CI 32.1—73.9), respectively. The probability of recovery conditional on not having died after one, two and three weeks of hospital admission was 7.6% (95% 5.6—10.3), 17.9% (95% CI 14.3—22.2).

As shown in figure 2, the cumulative incidence plot of death and recovery in the total population, separated by age group. Gray’s test indicated a significant difference between two groups for both death (p<0.01) and recovery (p <0.001). The probability of death for patients aged <50 years after one, two and three weeks of hospital admission was 4.6% (95% CI 1.5—13.8), 10.1% (95% CI 4.7—21.3), 12.6% (95% CI 6.1—24.9), respectively, whereas for patients aged >50 years and above, the probability of death was 30.9% (95% CI 19.9-46.0), 46.8% (95% CI 31.5—65.2), and 52.7% (95% CI 35.6-72.1), respectively.

Cumulative incidence plot of death and recovery in the total patients separated, by gender. Gray’s test indicated a statistically significant difference between both groups for death (p = 0.03), but not for recovery (p = 0.050). The probability of death for females after one, two and three weeks of hospital admission was 17.2% (95% CI 8.0—34.6), 21.1% (95% CI 10.5—39.8), and 47.4% (95% CI l6.4—90.0), respectively, whereas for males the probability of death was 18.7% (95% CI 11.4—29.8), 43.3% (95% CI 26.7-64.6), and 54.6% (95% CI 32.4-79.8), respectively.

As shown in figure 4, the cumulative incidence plot of death in the total patients separated, by type of co morbidities. Gray’s test indicated a statistically significant difference amongst the three groups for death (p = 0.03), but not for recovery (p = 0.16). The probability of death for hypertensive patients after one, two and three weeks of hospital admission was 26.2% (95% CI 10.7—64.4), and the same 46.2% (95% CI 17.2—1.24) after the rest two weeks, respectively, and for Immunocompromised patients the probability of death was 32.6% (95% CI 12.1—87.8), 52.6% (95% CI 20.0-1.38), and no death after the last week, respectively, whereas for diabetes mellitus patients the probability of death was the same 9.1% (95% CI 2.3—36.5) after all of the three consecutive weeks.

**Figure 4.**
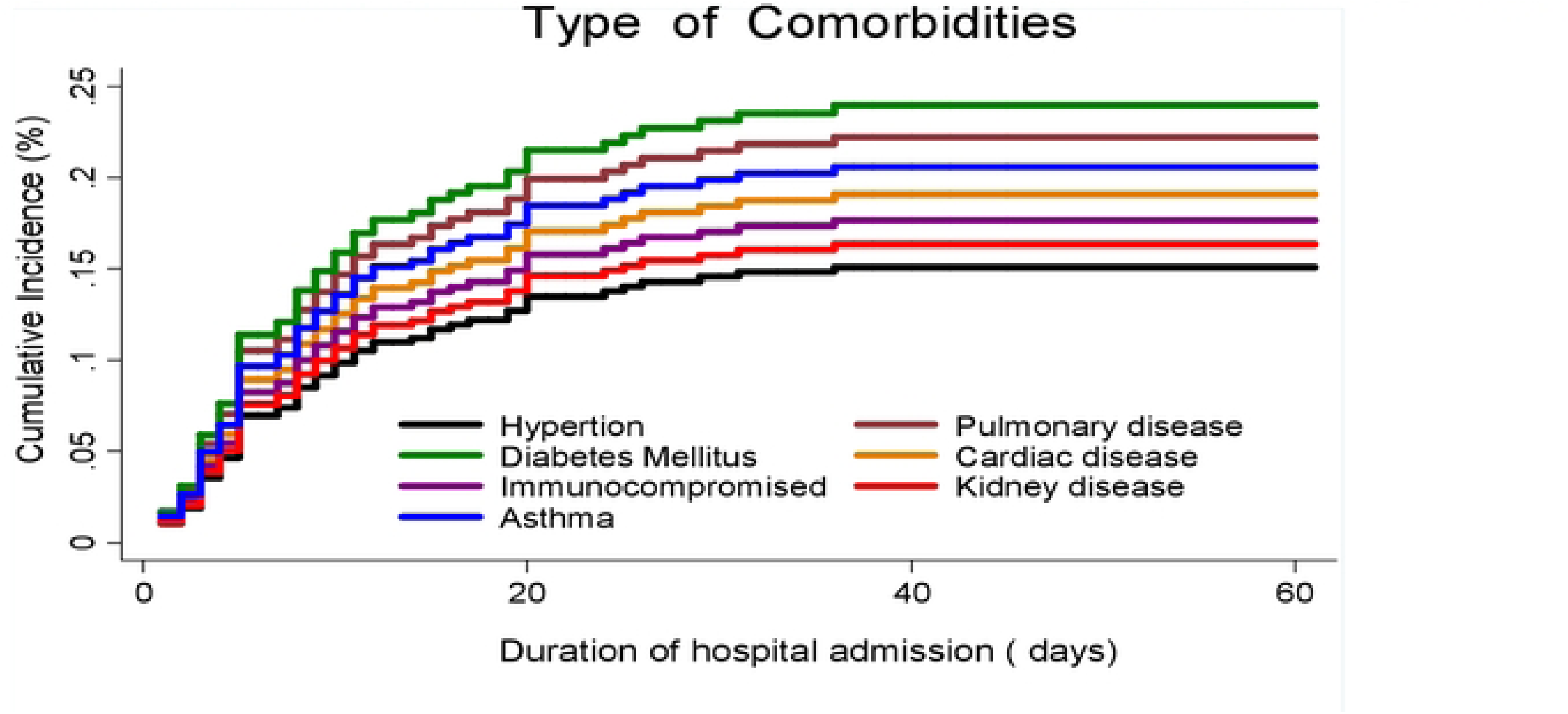
Cumulative incidence plots of death and recovery in south eastern Ethiopia hospitals, separated by type of Co morbidities 2024.

### Univariable and multivariable SH model

HRcs from univariable and multivariable SH models are reported in Table 2. Univariable analysis showed that older age increased the risk of death: with every year increase in age, the risk of death increased with 1% (HRcs 1.01, 95% CI 1.00—1.02), and also the chances of recovery insignificantly increased by 0.1% (HRcs 1.00, 95% CI 0.99—1.01). In terms of location, being in urban the risk of death increased with 90% (HRcs 1.90, 95% CI 1.16— 3.11), and also the chances of recovery increased by 29% (HRcs 1.29, 95% CI 1.04—1.59), regarding religion, being an Orthodox the risk of death increased with 196% (HRcs 2.96, 95% CI 1.50—5.82), and also the chances of recovery increased by 111% (HRcs 2.11, 95% CI 1.44—3.10), whereas, being a smoker the risk of death increased with 96% (HRcs 1.96, 95% CI 1.11—3.47), but the chance of recovery non significantly increased by 119% (HRcs 2.19, 95% CI 0.98—4.88). In terms of presenting symptom, symptomatic patients had an 97.4% increase in the risk of death (HRcs 1.974, 95% CI 1.29—3.01), but the chance of recovery decreased by 28% (HRcs 0.72, 95% CI 0.52—1.01)

**Table 2.**
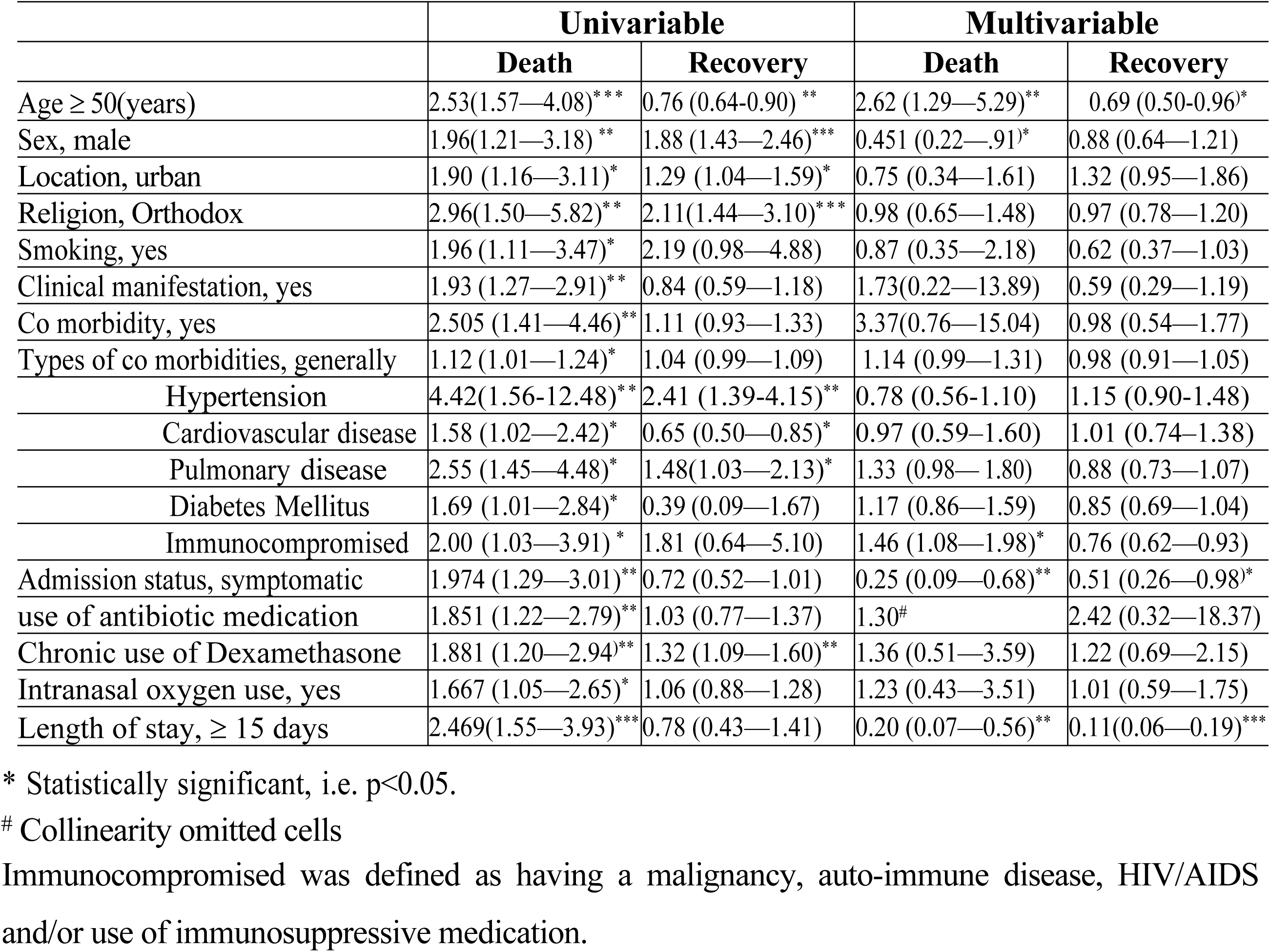
Univariable and multivariable cause-specific hazard ratios (HRcs) including 95% confidence intervals for death and recovery in south eastern Ethiopia hospitals, 2024.

|  | Univariable |  | Multivariable |  |
| --- | --- | --- | --- | --- |
|  | Death | Recovery | Death | Recovery |
| Age ≥ 50(years) | 2.53(1.57—4.08)*** | 0.76 (0.64-0.90) ** | 2.62 (1.29—5.29)** | 0.69 (0.50-0.96)* |
| Sex, male | 1.96(1.21—3.18) ** | 1.88 (1.43—2.46)*** | 0.451 (0.22—0.91)* | 0.88 (0.64—1.21) |
| Location, urban | 1.90 (1.16—3.11)* | 1.29 (1.04—1.59)* | 0.75 (0.34—1.61) | 1.32 (0.95—1.86) |
| Religion, Orthodox | 2.96(1.50—5.82)** | 2.11(1.44—3.10)*** | 0.98 (0.65—1.48) | 0.97 (0.78—1.20) |
| Smoking, yes | 1.96 (1.11—3.47)* | 2.19 (0.98—4.88) | 0.87 (0.35—2.18) | 0.62 (0.37—1.03) |
| Clinical manifestation, yes | 1.93 (1.27—2.91)** | 0.84 (0.59—1.18) | 1.73(0.22—13.89) | 0.59 (0.29—1.19) |
| Co morbidity, yes | 2.505 (1.41—4.46)** | 1.11 (0.93—1.33) | 3.37(0.76—15.04) | 0.98 (0.54—1.77) |
| Types of co morbidities, generally | 1.12 (1.01—1.24)* | 1.04 (0.99—1.09) | 1.14 (0.99—1.31) | 0.98 (0.91—1.05) |
| Hypertension | 4.42(1.56-12.48)** | 2.41 (1.39-4.15)** | 0.78 (0.56-1.10) | 1.15 (0.90-1.48) |
| Cardiovascular disease | 1.58 (1.02—2.42)* | 0.65 (0.50—0.85)* | 0.97 (0.59—1.60) | 1.01 (0.74—1.38) |
| Pulmonary disease | 2.55 (1.45—4.48)* | 1.48(1.03—2.13)* | 1.33 (0.98—1.80) | 0.88 (0.73—1.07) |
| Diabetes Mellitus | 1.69 (1.01—2.84)* | 0.39 (0.09—1.67) | 1.17 (0.86—1.59) | 0.85 (0.69—1.04) |
| Immunocompromised | 2.00 (1.03—3.91) * | 1.81 (0.64—5.10) | 1.46 (1.08—1.98)* | 0.76 (0.62—0.93) |
| Admission status, symptomatic | 1.974 (1.29—3.01)** | 0.72 (0.52—1.01) | 0.25 (0.09—0.68)** | 0.51 (0.26—0.98)* |
| use of antibiotic medication | 1.851 (1.22—2.79)** | 1.03 (0.77—1.37) | 1.30<sup>#</sup> | 2.42 (0.32—18.37) |
| Chronic use of Dexamethasone | 1.881 (1.20—2.94)** | 1.32 (1.09—1.60)** | 1.36 (0.51—3.59) | 1.22 (0.69—2.15) |
| Intranasal oxygen use, yes | 1.667 (1.05—2.65)* | 1.06 (0.88—1.28) | 1.23 (0.43—3.51) | 1.01 (0.59—1.75) |
| Length of stay, ≥ 15 days | 2.469(1.55—3.93)*** | 0.78 (0.43—1.41) | 0.20 (0.07—0.56)** | 0.11(0.06—0.19)*** |

\* Statistically significant, i.e. $p < 0.05$ .
<sup>#</sup> Collinearity omitted cells
Immunocompromised was defined as having a malignancy, auto-immune disease, HIV/AIDS and/or use of immunosuppressive medication.

Generally presence of co-morbidity increases the risk of death: Patients with two or more co morbidities had an 11.7% increase in the risk of death (HRcs 1.117, 95% CI 1.01—1.24), and also the chances of recovery increased by 3.6% (HRcs 1.036, 95% CI 0.98—1.08) but it was not statistical significant.

Of the co morbidities: Pulmonary disease, hypertension, HIV/AIDS and diabetes mellitus increases risk of death. Patients with pulmonary disease had a 155% increase in the risk of death (HRcs 2.55, 95% CI 1.45—4.84), and a 48% increase in chances of recovery (HRcs 1.48, 95% CI 1.03—2.13). Patients with hypertension had a 140.6% increase in the risk of death (HRcs 2.406, 95% CI 1.39—4.15), and a 341.6% increase in chances of recovery (HRcs 4.416, 95% CI 1.563—12.48). Whereas, HIV/AIDs Patients had a 100% increase in the risk of death (HRcs 2.002, 95% CI 1.03—3.91), and an 80.7% non significant increase in chances of recovery (HRcs 1.807, 95% CI 0.64—5.10). Furthermore, patients with diabetes mellitus had a 69% increase in the risk of death (HRcs 1.691, 95% CI 1.01—2.84), and a 39% non significant decrease in chances of recovery (HRcs 0.388, 95% CI 0.09—1.67).

In terms of medication, both use of antibiotics or dexamethason medication were associated with an increased risk of death (HRcs 1.85, 95% CI 1.22—2.79, and HRcs 1.88, 95% CI 1.20— 2.94, respectively). Regarding length of hospital stay and intranasal oxygen use or ventilatory support, both extended hospital stay and use of intranasal oxygen increased risk of death. Patients admitted ≥ 15 days had a 147% increase in risk of death (HRcs 2.47, 95% CI 1.55— 3.93) and patients with intranasal oxygen a 67% increase in risk of death (HRcs 1.67, 95% CI 1.05—2.65).

In multivariable analyses, the following factors were associated with risk of death: age in general, older age, gender, immunocompromised state, symptomatic state at time of admission, and extended length of stay after admission. Firstly, with every year increase in age, the risk of death increased with 2.3% (HRcs 1.023, 95% CI 1.00—1.05), and the chances of recovery increased with 0.3% (HRcs 1.003, 95% CI 0.993—1.014). In other words, if in two patients all variables except for age are the same, the patient who is one year older has a 2.3% higher risk of dying. Furthermore, patients with immunocompromised state had a 46% increased risk of death (HRcs 1.46, 95% CI 1.08— 1.98), and 24% decrease in chances of recovery (HRcs 0.76, 95% CI 0.62—0.93). Whereas, patients that were symptomatic at the time of admission had a 75% decrease in risk of death (HRcs 0.25, 95% CI 0.09—0.68), and a 49 % decrease in chances of recovery (HRcs 0.51, 95% CI 0.26— 0.98). Furthermore, aged patients ≥ 50 years at the time of diagnosis had a 162% increase in risk of death (HRcs 2.62, 95% CI 1.29—5.29). Finally, p a t i e n t s t h a t s t a y e d 1 5 o r m o r e d a y s following admission had an 80% decrease i n the risk of death (HRcs 0.20, 95% CI 0.07—0.56), and an 89% decrease in chances of recovery (HRcs 0.11, 95% CI 0.06—0.19).

## Discussion

### Summary of findings

In south eastern Ethiopia hospitals, approximately 17% of all COVID-19 patients died after a median hospital admission of five days during the hit of the epidemic. Using a competing risk approach, we identified age, gender, residence, status at admission, length of stay, co morbidities, such as pulmonary disease, hypertension, smoking, and use of medication as the most important risk factors in univariate analyses. After adjusting for all relevant factors at baseline, we found that higher age and immunocompromised state were associated with increased risk of death.

On the other hand, extended period of admission beyond 15 days was associated with lower mortality. Male sex was a significant protective factor about mortality.

### Interpretation of results and comparison to literature

Conventional survival analyses do not take competing risks into account, which leads to biased mortality estimates and to overestimation of survival curves. Using the competing risk approach, we took into account that patients who recovered were no longer at the same risk of dying than those who remained hospitalized, resulting in less biased mortality estimates. Even though death or recovery are the two possible final outcomes of the disease, the time to death and time to recovery may not the same (i.e. time to death was generally shorter than time to recovery). In addition, 21% of our patients were censored. As a result, the risk factors influencing death may differ from risk factors for recovery [30]. For example, we identified strong risk factors that both increased the risk of dying and reduced the recovery risk, namely higher age and immunosuppression. Male sex has reduced death risk in the final model, and symptomatic patients at admission and length of stay more than 15 days showed equivocal results, reducing the risk of death as well as the risk of recovery.

In our study population, approximately 1 7 % of all patients died, which is consistent with other studies [31,32]. Our study showed that age and low immune status at time of admission were risk factors for in-hospital mortality, which is in line with results of many other studies [18,33–35]. We included both cardiovascular disease and Asthma in our model, which may be subject to collinearity. However, a sensitivity analyses excluding cardiovascular disease from the model did not change our estimates and standard errors, showing that our reported estimates are valid. We found immunosuppression as a risk factor for mortality. This has been reported previously, although the exact role of the immune system in COVID-19 is complex [36–38].

Poor outcome could be determined by a declining immune system less able to clear the virus, but lung tissue damage in severe cases could also be caused by an exaggerated immune response, rather than damage inflicted by the virus itself [39–43]. Consequently, immunocompromised patients may be protected from this type of hyper inflammation [37,44,45]. We found that chronic use of antibiotics or dexamethason medication was associated with increased risk of death. Although this may be partially explained by the fact that these medications are used by patients with cardiovascular disease, which has been reported as an individual risk factor for in-hospital mortality [18], anticoagulant medication remained an independent risk factor for death in our multivariable analyses. Thromboembolic events are frequently reported in association with severe COVID-19 disease and mortality [46,47], and current guidelines suggest prophylactic anticoagulants in all hospitalized COVID-19 patients if not contraindicated. However, studies have reported conflicting results regarding the effect of anticoagulants medication on COVID-19 mortality [48], ranging from a protective effect [49] to a harmful effect [50,51], or no association [49,52]. Prospective studies and RCTs are needed to explore the true effects of these medications in hospitalized COVID-19 patients.

Findings of a living systematic review of 23 prognostic studies about COVID-19 mortality indicated that age, immunocompromising comorbidities, and composite scores of vital parameters are frequently reported predictors of in-hospital mortality, similar to our findings [18]. Blood ferritin levels and anticoagulants medication were scarcely reported [18]. On the other hand, male sex and comorbidities, such as cardiovascular and pulmonary diseases, that were significantly associated with in-hospital mortality in our univariable but not multivariable analyses, were frequently reported in other prognostic studies. This is an important finding and shows that many of these risk factors are interacting with each other.

Last, we found no increased risk of dying in patients with male sex, diabetes, which are also risk factors that have been reported previously. Larger studies may be needed to reveal additional risk factors of clinical importance.

### Strengths and limitations

The strength of this study is the large, multisite population in an area in which infections were clustered. We analyzed mortality in COVID-19 patients using a competing risk approach, leading to more accurate risk estimates for mortality than when using conventional survival analysis. As previously explained, conventional survival analyses may have resulted in biased estimates and Kaplan-Meier curves presenting overestimated incidence of death. There were some limitations to this study. First, approximately 21% of patients in our study were censored, mainly due to frequent transfer of patients. However, this was considered within acceptable limits [53–55]. Second, our data consisted of routinely collected data, resulting in missing values in several variables. We used multiple imputations to minimize associated bias and analyzing dropping missed values. The advantage of routinely collected data is that the study’s predictors are readily available and be used in clinical practice without extra effort. All reported predictors are variables measured at time of hospital admission, which means that these factors can be used to identify patients in need of more intensive care and monitoring in an early stage of hospitalization. Finally, data were collected in multiple centers with differences in manner of reporting in medical records and differences in clinical management. The advantage of this multisite approach is that it increased the generalizability of our findings.

## Conclusion

Using a robust, competing risk survival analysis, our study confirmed specific risk factors for in hospital COVID-19 mortality, adding rigor to the current knowledge of risk factors. We confirmed that age and immunocompromised state are important risk factors for death in hospitalized COVID- 19 patients. This allows us to corroborate specific pattern of risk factors about COVID-19 mortality and its progression among different groups of individuals (differentiated by age and immune-compromised state). Clinicians and administrators need to make arrangements to segregate and manage these individuals. Nearly 1 in 5 patients who succumb to illness in the hospitals are likely to do so within the first 72 hours. Further, those who die are likely to be older, with each advancing decade, conferring additional significant mortality risk. This should be considered while planning distribution of services. Nearly 1 of 2 patients who succumb to illness has associated immune-compromised state. Our findings reaffirm the need for prompt diagnosis, testing and multidisciplinary case mix management. Within health facilities these thoughts shall allow informed decision making for sound, effective, and sustained health-care services to ease mortality.

## Abbreviations

AHMC: Adama Hospital Medical College
AIDS: Acquired immunodeficiency syndrome
ARDS: Acute Respiratory Distress syndrome
AOR: Adjusted Odd Ratio
CI: Confidence Interval
COR: Crude Odd Ratio
HRcs: cause-specific hazard ratios
HIV: Human Immune Virus
ICU: Intensive Care Unit
IQR: Inter Quartile Range
KTC: Kurkura Treatment Center
PH: Primary Hospital
PPs: Proportional population size
OR: Odd Ratio
RH: Regional Hospital
RNA: Ribose Nucleotide Acid
SARS-COV-2: Severe Acute Respiratory Syndrome
SD: Standard Deviation
WHO: World Health Organization

## Declarations

### Ethical approval

This study was approved by Arsi University’s Health Research Ethical Review Committee, accreditation number ERC N<u>o</u> A/CHS/RC/75/2023 and permission was obtained from the treatment sites for the data collection. There was no direct human contact with the respondents in this study. Participant reports/information was already anonymized and de-identified prior to data collection. Data security and confidentiality were maintained at all levels of data management.

### Consent for publication

Not applicable

### Data Availability Statement

All relevant data are within the manuscript and the raw datasets used during the analysis were available from the corresponding author on reasonable request.

## Authors’ contributions

AW, AD, AKa, AT, MS, Aki, ZA and YM designed and worked on the study protocols. AD, AW and AT prepared a data collection tool and provided training to data collectors. AD and AT were conducted data entry to Epidata Entry client. AW, AD, AKa, AT, MS, Aki, ZA, and YM analyzed the data, interpreted the result, and wrote the manuscript’s draft and final version. All authors read and approved the final manuscript.

## Acknowledgments

Our heartfelt thanks go to Bishoftu, Modjo, Adama, Negele Arsi, Bekoji and Shashamane hospital administrations and staffs for their permission and cooperation during the data collection process.

